# Associations of Power Distance and Psychological Safety With Medical Researcher Well-being

**DOI:** 10.64898/2026.05.15.26353300

**Authors:** Jungmin Choi, Yeun Joon Kim, Yingyue Luna Luan

## Abstract

**OBJECTIVES:** To examine whether psychological safety and power distance are associated with medical researchers’ well-being, and whether these associations operate through team inclusiveness and conflict.

**DESIGN:** Cross-sectional survey study.

**SETTING:** A biomedical research institute at a major UK university.

**PARTICIPANTS:** 133 medical researchers from 17 teams, including 20 principal investigators and 113 team members.

**MAIN OUTCOME MEASURES:** Job satisfaction, life satisfaction, intrinsic motivation, and psychological detachment. Mediators were dimensions of team inclusiveness and team conflict.

**RESULTS:** Psychological safety had no significant direct associations with job satisfaction, life satisfaction, intrinsic motivation, or psychological detachment, but showed several indirect associations through researchers’ team experiences. It was indirectly associated with higher job satisfaction, life satisfaction, and intrinsic motivation primarily through greater integration of differences, inclusion in decision making, or more constructive forms of conflict (bs=.23-.38, ps=.032-<.001). For psychological detachment, psychological safety showed conflicting indirect associations: it had the potential to support detachment through greater integration of differences and lower avoidant conflict (bs=.21-.56, ps=.054-.002), but to undermine detachment through greater inclusion in decision-making (b=-.26, p=.082). Power distance showed a different pattern. Most notably, it was positively associated with psychological detachment (b=.54, p=.062). However, power distance was indirectly associated with lower job satisfaction, life satisfaction, and intrinsic motivation, primarily through reduced integration of differences and greater dominating conflict (bs=-.14 to -.19, ps=.068-.020).

**CONCLUSIONS:** Common assumptions about psychological safety and power distance should be revisited. Psychological safety did not show strong direct benefits for researcher well-being, whereas power distance was not uniformly harmful and was positively associated with psychological detachment. A more nuanced understanding of both cultural dimensions is needed in medical research teams.

## Introduction

Researcher well-being is fundamental to the progress and sustainability of medical science; when it deteriorates, the consequences extend beyond the individual, placing the quality and continuity of scientific work at risk.^1^ In an attempt to understand the drivers of researcher well-being, much of the existing discussion has centered on structural pressures, such as funding instability and high evaluative demands.^2^ While important, these factors are often outside the control of individual teams, offering limited guidance for improving researchers’ day-to-day experiences. More recent work has therefore begun to shift attention toward the immediate work environment as a critical, yet more actionable, determinant of well-being.^3^ In particular, team culture—defined as the shared beliefs, values, and assumptions that shape how members interact and work together^4 5^—plays a central role in structuring everyday experiences within research teams.^6^

Within medical research settings, two cultural dimensions are especially pivotal to researchers’ daily experiences: power distance and psychological safety. Medical research teams are typically organized around principal investigator (PI)-led hierarchies and operate under intense productivity pressures.^7 8^ These conditions often foster pronounced power distance, or the extent to which unequal distributions of power are accepted within a team,^9^ concentrating decision-making authority among senior members. Moreover, under high evaluative demands, psychological safety, or the shared belief that members can speak up, ask questions, or admit mistakes without fear of negative consequences,^10^ becomes a critical determinant of how junior researchers navigate professional risk. Together, these cultural dimensions directly capture how individuals navigate hierarchy and interpersonal risk in their daily work, making them particularly consequential for researchers’ experiences and well-being. The present research therefore investigates how power distance and psychological safety shape well-being among medical researchers.

In examining power distance and psychological safety, existing research has often employed a binary lens, portraying power distance as constraining and psychological safety as enabling key aspects of team functioning. In teams characterised by high power distance, members accept unequal distributions of power,^10^ which shapes how they engage with others in the team. Under this culture, lower-ranking members tend to prioritize deference to authority over expressing their own ideas, leading to reduced upward voice and limited participation.^11–13^ These dynamics are particularly pronounced in healthcare settings, where hierarchical structures are deeply embedded. High power distance may undermine collegiality by reinforcing rank over partnership and discouraging staff from engaging as equal peers.^14^ It may also increase fear of disclosing errors,^15^ further constraining open interaction.

In contrast to power distance, psychological safety has often been viewed as a facilitator of adaptive team functioning.^16^ Research across organizational settings suggests that psychological safety is associated with open communication, including voice, feedback seeking, and error reporting, as well as higher levels of learning and engagement.^10 17 18^ These patterns are also observed in healthcare contexts, where psychological safety has been linked to speaking up,^19^ reporting of adverse events,^20 21^ and a stronger sense of engagement and belonging among staff.^22^

Taken together, existing evidence suggests that power distance and psychological safety may differentially shape how researchers experience their work, which in turn may have implications for researchers’ well-being. In high power distance contexts, heightened concern about evaluation by senior members may limit individuals’ engagement and increase interpersonal strain—factors that have been associated with poorer well-being outcomes.^23 24^ In contrast, psychological safety has been linked to greater voice, feedback seeking, and open communication,^10^ which may foster a stronger sense of inclusion and engagement,^25^ which are associated with more positive well-being.^26 27^

However, the prevailing characterisation of power distance as detrimental and psychological safety as beneficial may be overly simplistic. While the risks associated with high power distance and the benefits of psychological safety are well documented, a more nuanced perspective suggests that both dimensions may involve trade-offs, simultaneously enabling and constraining different aspects of team functioning. For instance, although high power distance can suppress voice, it can also provide clearer role expectations and centralized responsibility, as principal investigators retain primary accountability for decisions and outcomes.^11^ This may reduce the extent to which team members feel personally responsible for potential mistakes or failures. In contrast, while psychological safety facilitates open communication and participation, it may also increase individuals’ sense of responsibility for their contributions,^28^ particularly when they are actively involved in discussions and decision-making. Such heightened involvement can create a form of psychological burden, making it more difficult to disengage from work during non-working hours.^29^

Despite these theoretical ambiguities, existing research offers an incomplete account of how power distance and psychological safety matter for medical researchers. Prior research has focused predominantly on functional outcomes that are closely tied to the constructs themselves, such as whether staff speak up, report errors, or engage in other safety-related behaviors.^15 19 20^ While these outcomes are important, this emphasis on observable behaviors provides only a partial view of how these dynamics operate in practice, with comparatively less attention to researchers’ psychological experiences. In particular, far less is known about how power distance and psychological safety shape the well-being of team members, despite its importance for sustaining the research workforce.

A further limitation concerns the empirical context of prior work. Existing evidence has largely been drawn from clinical and healthcare delivery settings, where the primary focus is on care processes and patient safety.^14 15 19 22^ In contrast, medical research teams have received comparatively little attention, despite their central role in generating the scientific knowledge that underpins medical innovation and ultimately shapes patient outcomes. Finally, prior work has paid limited attention to the mechanisms through which these cultural dimensions influence individual experiences. Without examining how power distance and psychological safety shape day-to-day experiences, it remains difficult to explain why these cultural characteristics may support some aspects of well-being while undermining others.

To address these limitations, the current research examines how power distance and psychological safety relate to medical researchers’ well-being. Following the mainstream paradigm in well-being research,^30^ we conceptualize well-being as a multidimensional construct, capturing not only individuals’ evaluative judgments about their work and life (job satisfaction and life satisfaction), but also their motivational experiences (intrinsic motivation) and their ability to disengage and recover from work (psychological detachment). This approach reflects the idea that well-being is not reducible to a single dimension, but instead encompasses how individuals feel about their work, how they engage with it, and how they regulate boundaries between work and non-work domains.

We further investigate whether these relationships are mediated by individuals’ everyday experiences within the team, focusing on experienced team inclusiveness and team conflict as key pathways through which cultural environments shape well-being. These experiences represent immediate psychological consequences of team culture, capturing how members experience their roles, interactions, and involvement in daily work. They are also conceptually linked to power distance and psychological safety: inclusiveness reflects the extent to which individuals feel involved and valued within hierarchical structures,^31^ and team conflict captures how disagreements are expressed and managed under conditions of greater or lesser interpersonal risk.^32^ Prior research has similarly connected these constructs to team culture, suggesting that they serve as critical mechanisms through which cultural environments are translated into individual experiences.^33 34^

## Methods

### Participants

An online survey was conducted with members of a specialized biomedical research institute at a major UK university, between July and September 2025. All current researchers within the institute were invited to participate. Invitations to participate in the survey were distributed via email to principal investigators (PIs) and their team members. A total of 20 principal investigators and 121 team members were invited to participate. We received 133 responses, including 20 from PIs and 113 from team members, representing 17 research teams. The response rate was 94.32%.

### Study Design

The survey was administered online using Qualtrics. Participants received an individualized survey link by email. After accessing the survey, they were provided with information about the study and gave informed consent before proceeding. All participants completed the survey independently, without discussion with other team members. Participants completed a questionnaire about their experiences within their current research teams. The survey included measures of power distance, psychological safety, experienced team inclusiveness, experienced team conflict, and individual well-being. Participants also provided demographic information, including age, gender, education, ethnicity, citizenship, and tenure at the institute. After completing the survey, participants were entered into a lottery for one of five £50 gift cards as compensation for participation. Figure 1 shows our conceptual model.

**Fig 1.**
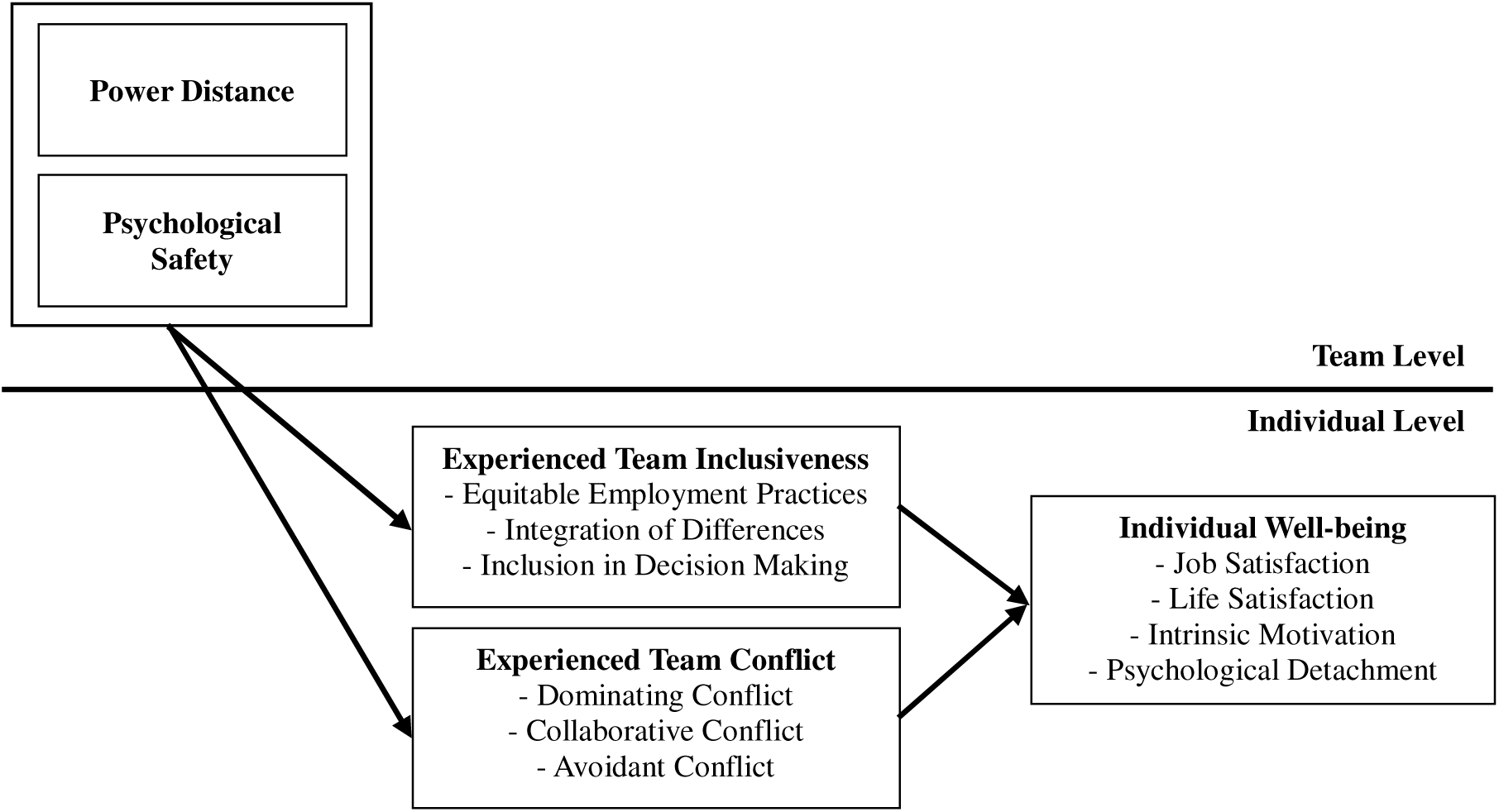
Conceptual model.

### Survey Instruments

Psychological safety and power distance were treated as team-level cultural variables. Psychological safety was assessed using a 7-item scale,^10^ with responses ranging from 1 (very inaccurate) to 7 (very accurate). Power distance was assessed using 5 items derived from the GLOBE study,^35^ rated on a 7-point scale. Consistent with the conceptualization of culture as a shared property of the team, individual responses were aggregated to the team level to capture collective perceptions of these cultural dimensions. Cronbach’s α was .88 for psychological safety and .80 for power distance.

Members’ experiences within their teams were assessed using measures of team inclusiveness and team conflict. Experienced team inclusiveness was measured using a 15-item instrument^31^ comprising three dimensions: equitable employment practices (5 items; α = .91), referring to fair and transparent procedures in allocating opportunities and resources; integration of differences (6 items; α = .88), reflecting openness to authenticity and diverse perspectives; and inclusion in decision making (4 items; α = .93), referring to the active involvement of members in team discussions and decision processes. Experienced team conflict was assessed using a measure of how disagreements are expressed and managed within teams,^32^ with three dimensions: collaborative conflict (4 items; α = .93), characterised by open dialogue and mutual problem-solving; dominating conflict (5 items; α = .87), reflecting the tendency to assert one’s position in order to prevail in conflicts; and avoidant conflict (4 items; α = .93), referring to suppressing or sidestepping disagreements. Responses for both constructs were recorded on a 7-point scale ranging from 1 (strongly disagree) to 7 (strongly agree).

Well-being outcomes included job satisfaction, life satisfaction, intrinsic motivation, and psychological detachment. Job satisfaction was assessed with three items from the Michigan Organizational Assessment Questionnaire (α = .95).^36^ Life satisfaction was measured using the five-item Satisfaction with Life Scale (α = .89).^37^ Intrinsic motivation was assessed with four items (α = .93).^38^ Psychological detachment from work during non-work time was measured using four items (α = .88).^39^ All of the outcomes were measured on a 7-point scale ranging from 1 (strongly disagree) to 7 (strongly agree). Full survey items are reported in the Supplementary Methods.

### Statistical Analysis

To examine whether team culture (i.e., psychological safety and power distance) influences individual well-being (i.e., job satisfaction, life satisfaction, intrinsic motivation, and psychological detachment) through experienced team inclusiveness and team conflict, we conducted a series of multilevel mediation analyses. Given the nested structure of the data—with individuals (Level 1) clustered within teams (Level 2)—all models were estimated using multilevel linear mixed-effects models via the lme4 package in R, specifying random intercepts for team membership.^40^

We tested two separate sets of mediation models. The first set examined experienced team inclusiveness as a mediator, operationalized across three parallel dimensions: equitable employment practices, integration of differences, and inclusion in decision-making. The second set examined experienced team conflict, similarly operationalized across three parallel dimensions: collaborative conflict, dominating conflict, and avoidant conflict. The two sets of mediators were tested in separate models rather than simultaneously, given their conceptual distinctiveness. All mediation analyses were conducted using the mediation package in R with 10,000 quasi-Bayesian Monte Carlo simulations.^41^ Mediation was inferred when the 95% confidence interval for the indirect effect excluded zero. There were no missing data for the variables included in the analyses.

## Results

The final sample (N = 133) consisted of 20 principal investigators (15.04%) and 113 team members (84.96%). The mean age of participants was 25.65 years (SD = 12.78). Of the sample, 79 participants identified as female (59.40%), 51 as male (38.35%), and 3 as non-binary or preferred not to disclose their gender (2.26%). In terms of ethnicity, 79 participants identified as White (59.40%), 39 as Asian/Pacific Islander (29.32%), 2 as Black (1.51%), 6 as Hispanic (4.51%), and 7 as Other or mixed race (5.26%). Regarding educational attainment, 69 participants held a PhD (51.88%), 30 a Master’s degree (22.56%), 25 a Bachelor’s degree (18.80%), 4 a professional degree (3.01%), 1 an associate degree (.75%), and 4 had completed some college or university without obtaining a degree (3.01%). The average tenure at the institute was 7.33 years (SD = 7.95).

### Psychological Safety and Well-Being

In our analyses, we distinguished between the direct effects of team culture and their indirect effects operating through the proposed mediators. For psychological safety, no significant direct associations were observed with any well-being outcomes, including job satisfaction (b = −.10, SE = .65, p = .649), life satisfaction (b = −.30, SE = .36, p = .397), intrinsic motivation (b = −.03, SE = .32, p = .936), and psychological detachment (b = −.37, SE = .04, p = .357).

Despite the absence of direct effects, psychological safety showed meaningful indirect associations with well-being through members’ experiences within the team (see Figure 2 and Table 1). Specifically, psychological safety was associated with higher job satisfaction through greater integration of differences, more collaborative forms of conflict, and lower levels of dominating conflict, with marginal evidence for an additional pathway through greater inclusion in decision-making. Similarly, psychological safety indirectly increased life satisfaction through greater integration of differences and, marginally, through higher collaborative conflict.

**Fig 2.**
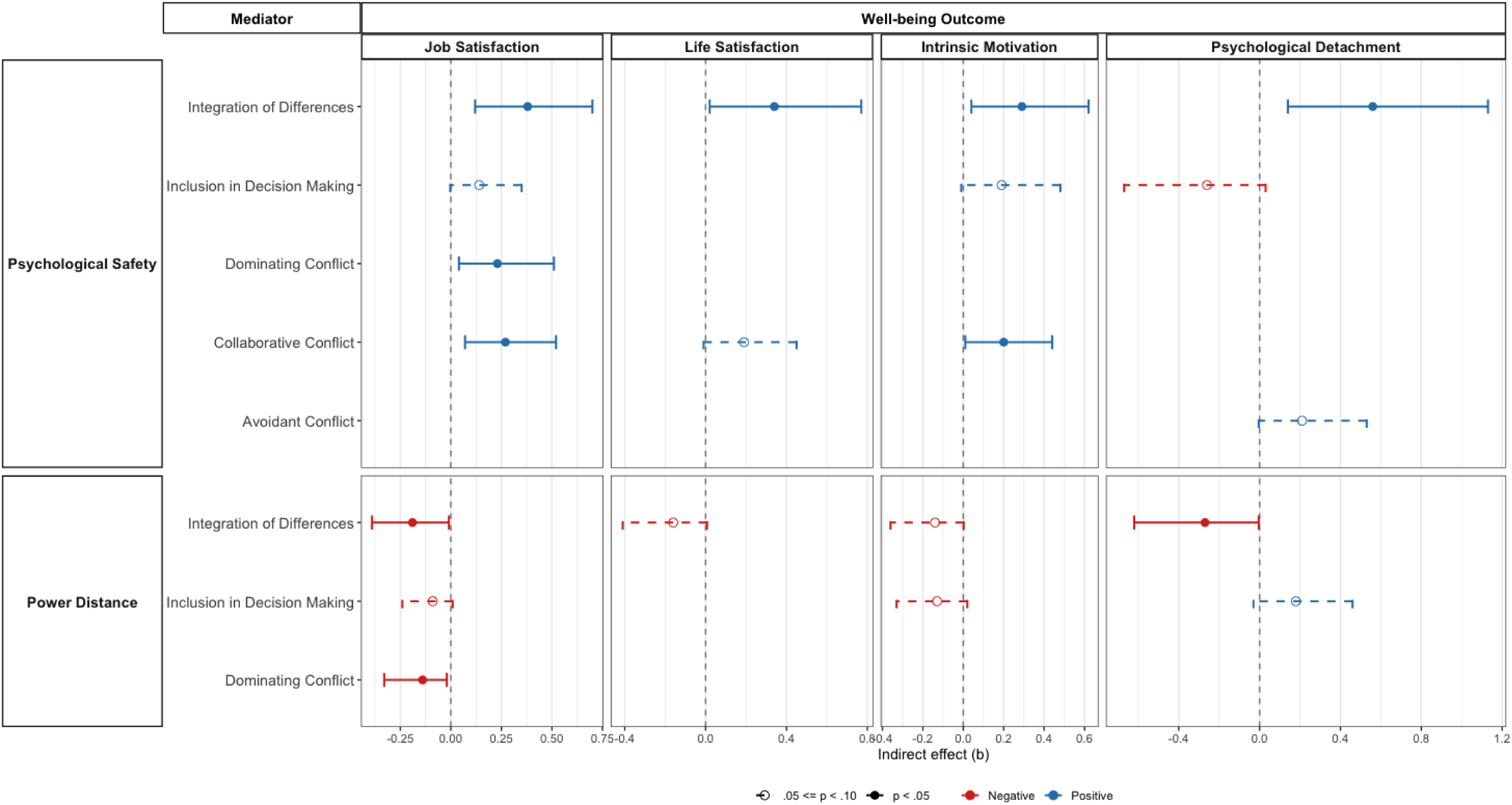
Forest plot of significant and marginal indirect effects of psychological safety and power distance on well-being through inclusiveness and conflict dimensions. Points represent indirect effect estimates (b), and horizontal lines denote 95% confidence intervals. Only statistically significant (p < .05) and marginally significant (.05 _≤_ p < .10) effects are displayed.

**Table 1.** Indirect effects of psychological safety and power distance on well-being via inclusiveness and conflict dimension.

| Culture | Mediator | Outcome |  |  |  |  |  |  |  |  |  |  |  |
| --- | --- | --- | --- | --- | --- | --- | --- | --- | --- | --- | --- | --- | --- |
|  |  | Job satisfaction |  |  | Life Satisfaction |  |  | Intrinsic Motivation |  |  | Psychological Detachment |  |  |
|  |  | *b* | *p* | 95% CI | *b* | *p* | 95% CI | *b* | *p* | 95% CI | *b* | *p* | 95% CI |
| Psychological Safety | Equitable Employment Practices | .05 | .408 | [-.07, .20] | .06 | .536 | [-.17, .33] | -.08 | .406 | [-.33, .09] | .01 | .894 | [-.27, .29] |
|  | Integration of Differences | .38 | < .001 | [.12, .70] | .34 | .022 | [.02, .77] | .29 | .012 | [.04, .62] | .56 | .002 | [.14, 1.13] |
|  | Inclusion in Decision Making | .14 | .062 | [ -.003, .35] | .04 | .704 | [-.17, .26] | .19 | .070 | [-.01, .48] | -.26 | .082 | [-.67, .03] |
|  | Dominating Conflict | .23 | .004 | [.04, .51] | .21 | .118 | [-.04, .58] | .07 | .538 | [-.16, .34] | -.08 | .590 | [-.44, .24] |
|  | Collaborative Conflict | .27 | .004 | [.07, .52] | .19 | .070 | [-.01, .45] | .20 | .032 | [.01, .44] | -.02 | .908 | [-.30, .21] |
|  | Avoidant Conflict | .05 | .356 | [-.05, .19] | .08 | .276 | [-.05, .32] | .06 | .334 | [-.05, .25] | .21 | .054 | [-.004, .53] |
| Power Distance | Equitable Employment Practices | -.06 | .212 | [-.18, .05] | -.09 | .346 | [-.31, .11] | .06 | .400 | [-.09, .25] | -.06 | .572 | [-.29, .17] |
|  | Integration of Differences | -.19 | .042 | [-.39, -.01] | -.16 | .068 | [-.41, .007] | -.14 | .058 | [-.36, .002] | -.27 | .046 | [-.62, -.004] |
|  | Inclusion in Decision Making | -.09 | .092 | [-.24, .009] | -.02 | .758 | [-.20, .12] | -.13 | .098 | [-.33, .02] | .18 | .090 | [-.03, .46] |
|  | Dominating Conflict | -.14 | .020 | [-.33, -.02] | -.12 | .146 | [-.35, .04] | -.04 | .556 | [-.21, .10] | .03 | .784 | [-.16, .24] |
|  | Collaborative Conflict | -.11 | .146 | [-.30, .04] | -.08 | .198 | [-.25, .03] | -.09 | .152 | [-.25, .03] | .002 | .962 | [-.11, .12] |
|  | Avoidant Conflict | -.02 | .454 | [-.11, .03] | -.05 | .322 | [-.20, .04] | -.04 | .400 | [-.15, .04] | -.01 | .150 | [-.34, .04] |

Psychological safety also indirectly increased intrinsic motivation through greater integration of differences and collaborative conflict, with marginal evidence supporting a pathway through greater inclusion in decision-making.

A more complex pattern emerged for psychological detachment, suggesting the presence of countervailing mechanisms. Psychological safety was associated with greater detachment through pathways such as increased integration of differences and, marginally, lower avoidant conflict. At the same time, it showed a marginal indirect association with lower detachment through greater inclusion in decision-making.

Overall, the findings indicate that psychological safety may shape well-being not through direct effects, but through its influence on members’ day-to-day experiences within the team. In particular, psychologically safe environments appear to support life satisfaction, job satisfaction, and motivation as they encourage openness to differing perspectives, inclusion in decision making, or constructive approaches to disagreement. Interestingly, psychological detachment appeared to be shaped by competing underlying processes. Although psychologically safe environments may facilitate disengagement from work through greater integration of differences and lower avoidant conflict, they may also foster stronger involvement in team interactions and decision-making, making it more difficult for members to mentally disconnect after work.

### Power Distance and Well-Being

Power distance showed no significant direct associations with job satisfaction (*b* = .17, SE = .15, *p* = .253), life satisfaction (*b* = .22, SE = .26, *p* = .390), or intrinsic motivation (*b* = -.01, SE = .22, *p* = .953). However, there was marginal evidence of a positive direct association with psychological detachment (b = .54, SE = .29, *p* = .062), indicating that greater hierarchical distance may “help” employees disengage from work.

As shown in Figure 2 and Table 1, power distance was linked to well-being primarily through indirect pathways operating via member experiences within the team. Specifically, power distance was negatively associated with job satisfaction through reduced integration of differences and greater dominating conflict, with marginal evidence for an additional pathway through lower inclusion in decision-making. Power distance was marginally negatively associated with life satisfaction indirectly through reduced integration of differences. Power distance also showed an indirect negative association with intrinsic motivation through reduced integration of differences, alongside marginal evidence for a pathway through lower inclusion in decision-making.

Although power distance showed a *positive* overall association with psychological detachment, the indirect pathways revealed a more complex pattern. Power distance was negatively associated with detachment through lower integration of differences, but marginally positively associated with detachment through lower inclusion in decision-making. These opposing pathways suggest that the positive overall association may be driven by additional, unmeasured mechanisms, such as clearer role boundaries, reduced decision responsibility, or lower cognitive involvement in team issues.

Taken together, these findings suggest that power distance may shape well-being primarily through members’ experiences within the team. Specifically, an environment characterised by higher power distance may lower job satisfaction, life satisfaction, and intrinsic motivation as it discourages the integration of differing perspectives, inclusion in decision-making, or fosters more dominating forms of conflict. However, surprisingly power distance was beneficial for psychological detachment.

### Discussion Principal Findings

The current research finds that psychological safety and power distance are associated with medical researchers’ well-being mainly through their experiences within the team.

Specifically, psychological safety was associated with higher job satisfaction, life satisfaction, and intrinsic motivation primarily through greater integration of differences, greater inclusion in decision-making, or more collaborative approaches to conflict. In contrast, higher power distance was associated with lower job satisfaction, life satisfaction, and intrinsic motivation through reduced integration of differences, lower inclusion in decision-making, or greater dominating conflict.

The findings for psychological detachment were more differentiated. Psychological safety had only indirect and conflicting associations with psychological detachment, whereas power distance showed a positive overall association with detachment. Specifically, psychologically safe environments had the potential to support greater psychological detachment through higher integration of differences and lower avoidant conflict, suggesting that open engagement with differing perspectives may help researchers disengage from work during non-work time. At the same time, psychological safety also had the potential to undermine psychological detachment through greater inclusion in decision-making, suggesting that deeper involvement in team discussions and decision processes may extend cognitive and emotional engagement with work beyond formal working hours. These opposing pathways may explain why the overall association between psychological safety and detachment was non-significant. By contrast, although power distance was positively associated with psychological detachment overall, the mechanisms examined in this study do not appear to fully explain this association. We suggest that role clarity may be one mechanism explaining this positive association. In more hierarchical teams, decision-making authority and responsibility may be more clearly concentrated among senior members, which could reduce junior researchers’ sense of responsibility for unresolved team issues and make it easier for them to mentally disconnect from work after hours.

### Relation to Previous Research

The current research extends previous literature in several ways. First, the findings challenge the prevailing binary characterisation of psychological safety as enabling and power distance as constraining. Contrary to much prior research, psychological safety did not show a significant direct association with well-being outcomes; its associations were observed mainly through indirect, and in some cases relatively weak, pathways via researchers’ team experiences. By contrast, power distance showed a significantly positive direct association with psychological detachment, suggesting that hierarchical team cultures may not be uniformly detrimental. These findings point to a more differentiated view of team culture, in which psychological safety may support well-being only through specific interpersonal mechanisms, while power distance may also have unexpected benefits for recovery from work. More broadly, the results show the importance of distinguishing among different dimensions of well-being rather than assuming that cultural conditions considered desirable in one respect will produce uniformly positive outcomes.

Second, the current research suggests that the associations between team culture and well-being are shaped primarily through researchers’ day-to-day experiences within the team. By highlighting the role of mechanisms such as the integration of differences, inclusion in decision-making, and different forms of conflict, the findings clarify how broader cultural environments are translated into lived experiences that ultimately shape individual well-being. In particular, the integration of differences emerged as a central pathway linking team culture to well-being, suggesting that the extent to which research teams enable members to express and work through differing perspectives may be especially important for sustaining well-being in collaborative scientific work. The findings also point to inclusion in decision-making as a more differentiated mechanism. Although participatory practices are typically regarded as beneficial, greater involvement in team decision-making may also intensify individuals’ cognitive and emotional engagement with work, making disengagement and recovery more difficult.

The current research also contributes to previous literature by extending the study of psychological safety and power distance beyond behavioral and safety-related outcomes that have traditionally dominated this area of research.^15 19 20^ Whereas prior work has focused primarily on outcomes such as speaking up, error reporting, and other observable workplace behaviors, the present findings draw attention to how these cultural dimensions relate to researchers’ psychological well-being. In doing so, the study broadens understanding of how psychological safety and power distance shape researchers’ experiences within medical research teams, suggesting that their implications extend beyond communication and safety behaviors to encompass motivation, satisfaction, and recovery.

In addition, the present study addresses an important gap in the empirical context of prior research. Existing evidence on psychological safety and power distance has been derived predominantly from clinical and healthcare delivery settings, where concerns surrounding patient care and safety are central.^14 15 19 22^ By focusing on medical research teams, the current findings extend this literature to a context characterised by intensive collaboration, dependence on senior investigators, and strong evaluative pressures.

### Practical Implications

The results offer important implications for PIs and leaders of medical research teams.

Most importantly, they suggest that leaders should avoid treating psychological safety as universally beneficial or power distance as uniformly harmful. Although psychological safety is often promoted as an unquestionably positive feature of team culture, our findings suggest a more cautious view. Psychological safety may support researcher well-being by encouraging openness and constructive engagement, but its effects were largely indirect and relatively weak. Leaders should therefore foster psychological safety not as an end in itself, but as a condition that enables more specific beneficial experiences, such as the integration of different perspectives and constructive forms of conflict. At the same time, they should recognise that highly participatory environments may also increase researchers’ cognitive involvement in team issues, making it harder for them to detach from work.

The findings also suggest that some features of power distance may have value when managed appropriately. Although some indirect pathways linked power distance to lower satisfaction and motivation, power distance was directly and positively associated with psychological detachment. One possible explanation is that hierarchical cultures provide clearer role expectations, decision boundaries, and lines of responsibility, which may help researchers disengage from work during non-work time. The practical challenge for leaders, therefore, is not simply to eliminate power distance, but to manage hierarchy in a way that preserves clarity without suppressing voice. PIs may need to provide clear authority structures and responsibility boundaries while still ensuring that junior researchers can raise concerns, contribute ideas, and have their perspectives taken seriously. In this sense, effective leadership in medical research teams requires a careful balance: enough psychological safety to support openness and learning, but enough role clarity to prevent participation from becoming an unbounded burden.

### Limitations and Future Research

This study has several limitations. First, the cross-sectional design limits causal inference regarding the relationships between psychological safety, power distance, team experiences, and well-being. Future longitudinal research would help clarify how these cultural dimensions shape researchers’ well-being over time and whether these relationships operate reciprocally.

Second, the findings for psychological detachment suggest that the mechanisms examined in the present study may capture only part of the relationship between power distance and disengagement from work. Although higher power distance was associated with greater psychological detachment overall, the indirect pathways operated in opposing directions. Future research may therefore benefit from examining additional mechanisms through which power distance shapes disengagement and recovery from work.

Third, the present findings were based on data collected from researchers within a single medical research institute, which may limit the generalisability of the findings to other research environments. Team structures, leadership practices, and norms may differ across institutions and national contexts. Future research may therefore examine whether similar patterns emerge across different types of medical research organisations and research settings.

Finally, future research may further investigate contextual factors that shape when psychological safety and power distance are most likely to support or undermine well-being. For example, the consequences of psychological safety may differ depending on workload, role clarity, or preferences for autonomy within the team. Similarly, the implications of power distance may vary under conditions of time pressure, uncertainty, or resource scarcity. Examining such boundary conditions may help clarify when psychologically safe or hierarchical team environments are most conducive to sustainable well-being.

## Data Availability

Deidentified data are not available for sharing due to the small number of teams and the sensitive nature of the workplace data, which may pose a risk of participant identification.

## Ethics approval

Ethics Committee of the University of Cambridge gave ethical approval for this work (approval no. 24-30).

