## Supplemental Methods for "Associations of Power Distance and Psychological Safety With Medical Researcher Well-being"

### **Supplementary Methods**

#### **Survey Items**

**Psychological Safety (1 = Very inaccurate, 7 = Very accurate)**

1. Even if you make a mistake on my team, it is not held against you.
2. Members of my team are able to bring up problems and tough issues.
3. People on my team accept others, even when they are different.
4. It is safe to take risks on my team.
5. It is easy to ask other members of my team for help.
6. No one on my team would deliberately act in a way that undermines my efforts.
7. When working with members of my team, my unique skills and talents are recognized, valued, and utilized.

**Power Distance**

1. In my team, a member's influence is based primarily on (one’s ability and contribution to the team: 1; the authority of one’s position: 7).
2. In my team, members are expected to (obey the PI without question: 1; question the PI when in disagreement: 7).
3. In my team, members in positions of power try to (increase their social distance from less powerful members: 1; decrease their social distance from less powerful members: 7).
4. In my team, rank and position in the hierarchy come with special privileges (strongly agree: 1; strongly disagree: 7).
5. In my team, power is (concentrated at the top: 1; shared throughout the team: 7).

**Experienced Team Inclusiveness (1 = Strongly disagree, 7 = Strongly agree)**

*Equitable Employment Practices*

1. My team has a fair process (e.g., when assigning projects and determining authorship).
2. The performance review process (e.g., when determining authorship) in my team is fair.
3. My team invests in the development of all its members.
4. Members of my team receive equal rewards and compensation for equal work.
5. My team provides safe ways for members to voice their grievances.

*Integration of Differences*

1. My team is characterized by a non-threatening environment where members can reveal their true selves.
2. My team values work-life balance.
3. My team commits resources to ensuring that members can resolve conflicts effectively.
4. Members of my team are valued for who they are as people, not just for their roles.
5. In my team, members often share and learn about one another.
6. My team has a culture in which members appreciate the differences that individuals bring to the team.

*Inclusion in Decision Making*

1. In my team, members' input is actively sought.
2. In my team, everyone’s ideas for improvement are given serious consideration.
3. In my team, members' insights are used to rethink or redefine work practices.
4. Team leadership believes that problem-solving improves when input from different roles and perspectives is considered.

**Experienced Team Conflict** **(1 = Strongly disagree, 7 = Strongly agree)**

*Dominating Conflict*

1. Team members push their own points of view
2. Each team member searches for gains only for themselves
3. Team members fight for what they personally want.
4. Team members do everything to win for themselves
5. Team members try to force others to accept their points of view

*Collaborative Conflict*

1. Team members examine issues until they find a solution that satisfies everyone
2. Team members examine ideas from all sides to find a mutually optimal solution
3. Team members work out a solution that serves everyone’s interests.
4. Team members try to come up with creative solutions that incorporate multiple perspectives.

*Avoidant Conflict*

1. Team members avoid discussing conflict openly.
2. Team members avoid openly discussing conflict.
3. Team members are very reluctant to talk openly about conflict.
4. Conflict is not dealt with openly in my team.

**Individual Well-being (1 = Strongly disagree, 7 = Strongly agree)**

*Job Satisfaction*

1. Generally speaking, I like working with my team
2. In general, I like working with my team
3. In general, I like my job with my team

*Life Satisfaction*

1. In most ways, my life is close to my ideal.
2. The conditions of my life are excellent.
3. I am satisfied with my life.
4. So far, I have gotten the important things I want in life
5. If I could live my life over, I would change almost nothing.

*Intrinsic Motivation*

1. I like my job.
2. My job is fun.
3. I find my job engaging.
4. I enjoy my job.

*Psychological Detachment*

After work hours,

1. I forget about work.
2. I don’t think about work at all.
3. I distance myself from my work.
4. I get a break from the demands of work.
